# Development and Validation of an MGBP Nomogram for 3-month Unfavorable Functional Prognosis Prediction of Nontraumatic Pediatric Intracerebral Hemorrhage in Chinese Patients: 10-year Experience from a National Children’s Medical Center

**DOI:** 10.1101/2023.02.03.23285464

**Authors:** Ji-Hua Zhou, Zhen-Yu Zhang, Yang Chen, Mei-Xiu Ming, Quan-Li Shen, Ji-Cui Zheng, Gang Pan, Yi Zhang, Wei-Ming Chen, Guo-Ping Lu

## Abstract

**Background:** Pediatric intracerebral hemorrhage (pICH) remains a considerable cause of mortality. This study aimed to explore functional prognosis predictors in Chinese patients with pICH and attempted to develop and validate a nomogram for estimating individual risk probability of short-term unfavorable functional prognosis.

**Methods:** A retrospective case-control study through records reviewing was conducted, non-neonate patients with nontraumatic pICH discharged from the Children’s Hospital of Fudan University between January 2012 and December 2021 were all originally included. The primary outcome was unfavorable functional prognosis at 3 months post pICH defined as a score of 3–6 as measured using the modified Rankin Scale (ranging from 0 [asymptomatic] to 6 [death]). Multivariate logistic regression analysis was performed to screen prognosis predictors, a prediction model-based nomogram was developed. Internal validation was assessed and quantified as receiver operating characteristic (ROC) curve and bootstrapped calibration curve.

**Results:** A total of 269 pICH patients were enrolled, the median age was 57.2 months (interquartile range, 5.4–115.1), and 157 (58.4%) patients were male. The median follow-up time was 3.2 months (interquartile range, 2.8–3.6), 112 (41.6%) patients had unfavorable functional outcome. Cerebral vascular *m*alformation as etiology, modified *G*lasgow Coma Score on admission, *b*rainstem location, and intracerebral hemorrhage volume as *p*ercentage of total brain volume were identified as functional prognosis predictors. A nomogram was established comprising these four predictors, abbreviated to MGBP, the area under the ROC curve of the nomogram was 0.827 (95% Confidence Interval, 0.778–0.877) with good calibration (*P*=0.803 for the Spiegelhalter’s Z-test).

**Conclusions:** The MGBP nomogram is the first model developed and validated in a consecutive cohort to predict 3-month unfavorable functional prognosis post nontraumatic pICH in Chinese children, which may provide clinicians with a potentially effective approach for early prediction and timely management of pICH.

## Introduction

Intracerebral hemorrhage (ICH) is defined as brain injury due to acute blood extravasation into the brain parenchyma from a ruptured cerebral blood vessel.^1^ Nontraumatic hemorrhagic stroke (HS), including ICH, intraventricular hemorrhage (IVH), and subarachnoid hemorrhage (SAH) accounts for approximately half of non-neonate pediatric stroke, with annual incidence of approximately 1.0–1.7 per 100000.^2-4^ Pediatric ICH (pICH) represents approximately one-fourth to two-thirds of pediatric HS.^3^

As reported from a recent meta-analysis across 19 studies, although pICH is a rare event, aggregate case-fatality rate was 17.3% at last follow-up. Moreover, the complete recovery rate was only 27%, functional deficits were documented in a large proportion of survivors.^5^ Early prediction of unfavorable prognosis undoubtedly aids clinicians in precise patients’ management and helps patients and parents understand the medical consequences. Previous studies, mostly from North America or Europe, reported scattered data, comprehensive screening of prognosis predictors based on multivariate analysis was rarely performed due to small sample size,^6-9^ a few indicators including etiology, altered mental status, location, ICH volume, IVH presence, brain herniation, and hydrocephalus may have potential association with poor functional outcome post pICH.

However, research regarding pICH in Chinese children remain scant, prior literature mainly focus on case-series reports,^10-15^ whether the prognostic factors in Chinese pICH cohort are similar to those described for other ethnic is unknown, therefore, predicting the risk probability of unfavorable functional prognosis would be meaningful to support clinical decision making.

Multi-parametric nomogram is a graphical calculation tool which calculates the probability of an event based on individual characteristics with better visualization, as a novel prediction approach it has been commonly applied within adult stroke practice to improve predictive accuracy.^16-18^ Accordingly, we aimed to identify predictors of functional outcome in Chinese patients with nontraumatic acute pICH and to construct a simple visualized nomogram approach for individualized prediction of short-term unfavorable functional prognosis.

## Methods

### Data Availability

The data sets that support the findings of this study are available from the corresponding author on reasonable request. The corresponding author takes responsibility for data integrity and the data analysis.

### Study Protocol Approval and Patient Consents

The Institutional Review Board of Children’s Hospital of Fudan University (CHFU, a National Children’s Medical Center, Shanghai, China) approved the study protocol. Informed consent was waived because of electronic records in this study database were deidentified and the nature of this retrospective study.

### Study Population, Follow-up and Outcome Measurement

A case-control based prediction model study was conducted through code search (International Classification of Diseases, 10^th^ Revision [ICD-10] code I61) from the database of CHFU to identify nontraumatic, spontaneous ICH from all hospitalized non-neonate pediatric patients who were discharged between 1st January 2012 and 31st December 2021, cases with consecutive nontraumatic pICH were confirmed through medical records reviewing, relevant data including demographics, clinical and radiological features and functional outcomes of pICH were retrospectively extracted and recorded for analyses.

Inclusion criteria were as follows: (1) children aged 29 days to 18 years; (2) nontraumatic acute intraparenchymal hemorrhage (IPH) with radiological confirmation through head neuroimaging (computed tomography or magnetic resonance image) and admitted within 72 hours post initial symptom onset; (3) first-ever stroke. Exclusion criteria were as follows: (1) pre-term newborns whose adjusted gestational age on admission was ≤ 41 weeks; (2) isolated IVH or SAH without IPH; (3) intracranial tumor; (4) hemorrhagic transformation of arterial ischemic stroke (AIS) or cerebral sinovenous thrombosis; (5) incomplete medical records or neuroimaging could not be abstracted.

Short-term neurological follow-up was scheduled for every survivor at 90 days (7 days before or after were acceptable) post initial symptom onset. Through face-to-face clinic visit, functional impairment was evaluated by a multidisciplinary team comprising pediatric neurosurgeon, neurologist, neurointensivist and rehabilitation specialist using standard protocol with a structured interview, residual symptoms and functional deficits were recorded.

The primary outcome measure was 3-month unfavorable functional prognosis. Since no gold-standard pediatric stroke outcome assessment exists, the modified Rankin Scale (mRS) score was retrospectively graded by two investigators independently (one senior pediatric neurosurgery [J.C.Z.] and one senior pediatric neurologist [G.P.], both were blinded to hospitalization records and neuroimaging data) through follow-up records reviewing, discrepancies were reviewed until a consensus was reached. The mRS, ranging from 0 (asymptomatic) to 6 (death), is a validated scale widely applied for evaluating stroke patient outcomes.^16,19-24^ Favorable and unfavorable functional prognosis were defined by mRS score of 0–2 (asymptomatic to slight disability) and 3–6 (moderate disability to death), respectively, at 3 months post pICH.

### Definitions and Measurements of Potential Predictors

In accordance with our institutional protocol of pICH, mental status on admission was assessed through modified Glasgow Coma Score (GCS) ranging 3–15. Noninvasive systolic blood pressure (SBP) and diastolic blood pressure (DBP) on admission were measured three times and values were averaged. Based on 2017 guideline of high blood pressure (BP) in children and adolescents,^25^ for children aged 1–13 years, hypertension was classified as SBP and/or DBP≥95^th^ percentile or 130/80 mmHg (whichever is lower) by sex, age and height; for children aged≥13 years, hypertension was classified as≥130/80 mmHg; and according to Report of the Second Task Force on BP Control in Children--1987,^26^ for infants aged 29–30 days and 31 days–<1 year, hypertension was classified as SBP≥104 mmHg and≥ 112/74 mmHg, respectively. According to the recommendations for glucose management from 2022 guideline of spontaneous ICH (sICH),^1^ hyperglycemia was defined as serum glucose concentration higher than 10.0 mmol/L (180mg/dL); hypoglycemia was defined as serum glucose concentration of 3.3 mmol/L (60mg/dL) or lower. Admission modified GCS, BP and serum glucose concentration were measured within the first hour of hospitalization from admission. Etiologies of pICH were confirmed by medical history, laboratory tests, neuroimaging, results from surgical operation, interventional therapy and pathological biopsy. Hospitalization records were reviewed for demographic and clinical features extraction by a senior pediatric intensivist [Y.C.] who was blinded to neuroimaging data and follow-up records.

ICH volume (ICHV) and total brain volume (TBV) were estimated separately with the simple formula ABC/2 and XYZ/2,^27^ and ICHV was shown as percentage of TBV with the formula ABC/XYZ previously modeled to be an accurate reflection of hemorrhage size in pICH,^28^ an example of hemorrhage measurement is illustrated in Figure S1. Neuroimaging data were reviewed for hemorrhage features extraction and ICHV/TBV was measured retrospectively through manual tracing taken by two separate investigators and averaged (one senior pediatric radiologist [Q.L.S.] and one senior pediatric neurointensivist [Z.Y.Z.], both were blinded to clinical data and initial radiographic reports).

### Statistical Analysis

As it was a multivariate analysis, rules of thumb were adopted for sample size estimation, that is, the sample size should be 10 times the number of variables. Based on literature review, we proposed 10–20 variables as potential predictors, therefore, a sample size of 100–200 would meet the requirement.^29^

Statistical Package for the Social Sciences software (version 26.0, IBM SPSS Statistics, Chicago, IL, USA) was used for descriptive statistics, logistic regression model constructing and equation fitting. Continuous variables were reported as median and interquartile range (IQR) values. Categorical variables were expressed as proportions. A two-sided *P*<0.05 was defined as statistical significance.

Univariate and multivariate binary logistic regression analyses were performed to identify prognosis predictors. Statistically significant variables (*P* < 0.1) in univariate analysis were further included in multivariate analysis, using a stepwise regression (Likelihood ratio tests of Maximum Likelihood estimation) to determine the variables that would eventually enter the regression equation, regression coefficients, odds ratio (OR) with 95% confidence intervals (CI) for each of the significant prognosis predictors were calculated for logistic regression model establishment. Model visualization, nomogram development and validation, and performance evaluation were performed with R statistical software (version 4.0.1, R Foundation for Statistical Computing, Vienna, Austria) through discrimination statistics, resampling techniques (bootstrapping for internal validation), calibration statistics and potential clinical utility analysis. In this study, a prediction model was constructed using complete-case analysis only. Data analyses were conducted between June to September 2022. The TRIPOD guideline for reporting prediction studies was followed in drafting manuscript.

## Results

### Demographics, Clinical and Hemorrhage Characteristics

From the electronic database of CHFU, we assessed 366977 non-neonate pediatric in-patients who were discharged between January 2012 to December 2021 and identified 314 nontraumatic acute ICH cases through ICD-10 code search, 298 cases were confirmed nontraumatic acute ICH in non-neonate pediatric patients through medical records reviewing. Overall incidence of nontraumatic acute pICH in hospitalized non-neonate patients was 0.81‰ (298/366977) at our institution over the past decade, suggesting an extremely rarity of pICH, and we did not observe any identifiable trend by study period. Detailed incidence rate of each year is presented in Table S1.

29 cases among 298 confirmed nontraumatic acute ICH patients met the exclusion criteria, a total of 269 patients were finally enrolled in our study, which was able to meet the sample size requirement, all enrolled cases were utilized without missing values, a detailed flowchart of patients’ selection is displayed in Figure 1. None of the patients had history of preexisting diabetes or pre-morbid baseline neurological dysfunction.

**Figure 1.**
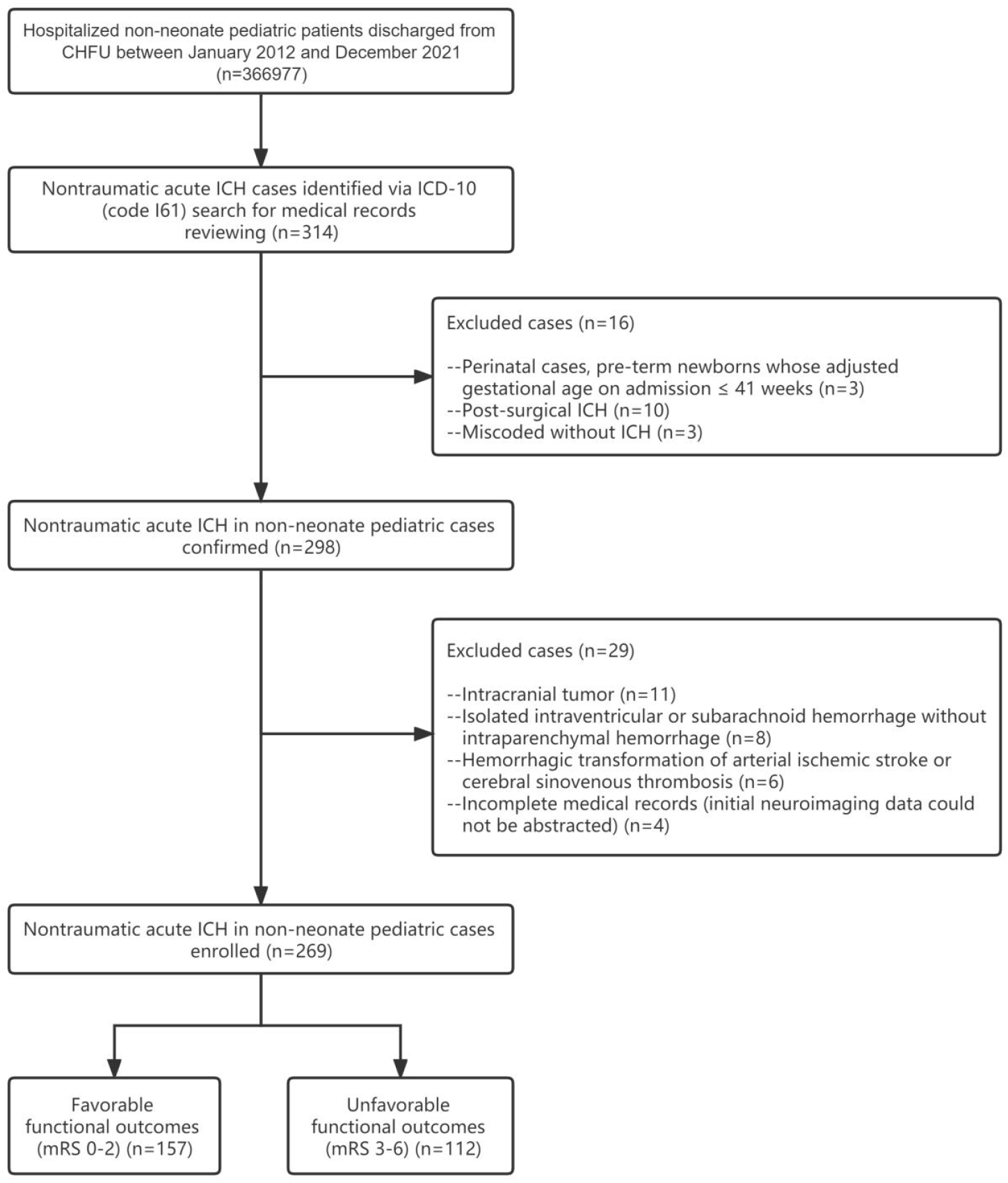
Flowchart for case identification. CHFU indicates Children’s Hospital of Fudan University; ICH, Intracerebral hemorrhage; and mRS, modified Rankin Scale.

Demographics, clinical and hemorrhage characteristics of the pICH cohort are described in Table 1. Median (Interquartile range, IQR) age on admission was 57.2 (IQR, 5.4–115.1) months, it has a bimodal distribution with the highest incidence in infancy (89, 33.1%) and the second highest incidence in early adolescent (46, 17.1%). 157 (58.4%) cases were male and male-to-female ratio was 1.4:1. Cerebral vascular malformation (113, 42.0%) was the dominant etiology category observed in this study, with arteriovenous malformation (AVM) as the most frequent in 82 (30.5%) patients, followed by cavernous malformation in 27 (10.0%), up to 72.6% and 23.9% of all cerebral vascular malformations, respectively. Hemostatic disorders associated ICH (HDICH) (84, 31.2%) were in the second place, idiopathic vitamin K deficiency bleeding (VKDB) in 29 (10.8%) ranked first (accounted for 34.5% of all HDICH), yet hemophillia in 3 (1.1%) was not common. As the third largest etiology category, 29 (10.8%) cases remained idiopathic (unknown etiology) despite thorough investigations during acute hospitalization and subsequent follow-up. Intracranial infection (27, 10.0%) and systemic diseases (16, 5.9%) were less common, specific etiological spectrum are summarized in Table S2. Etiology varied by age, HDICH represented over half of lesions in infantile subgroup; meanwhile, in all the remaining age subgroups ranging 1–18 years, vascular malformation represented over half of lesions in each subgroup, indicating age is a key clue in the diagnostic workup, detailed distribution of etiology categories with age subgroup analysis are shown in Figure S2. Initial clinical presentation also varied by age, details in the entire cohort and in infantile subgroup are demonstrated in Table S3 and Table S4. Of 269 cases with pICH, 73.2% were admitted to pediatric intensive care unit (PICU), 21.2% were intubated, indicating high critical care utilization.

**Table 1.**
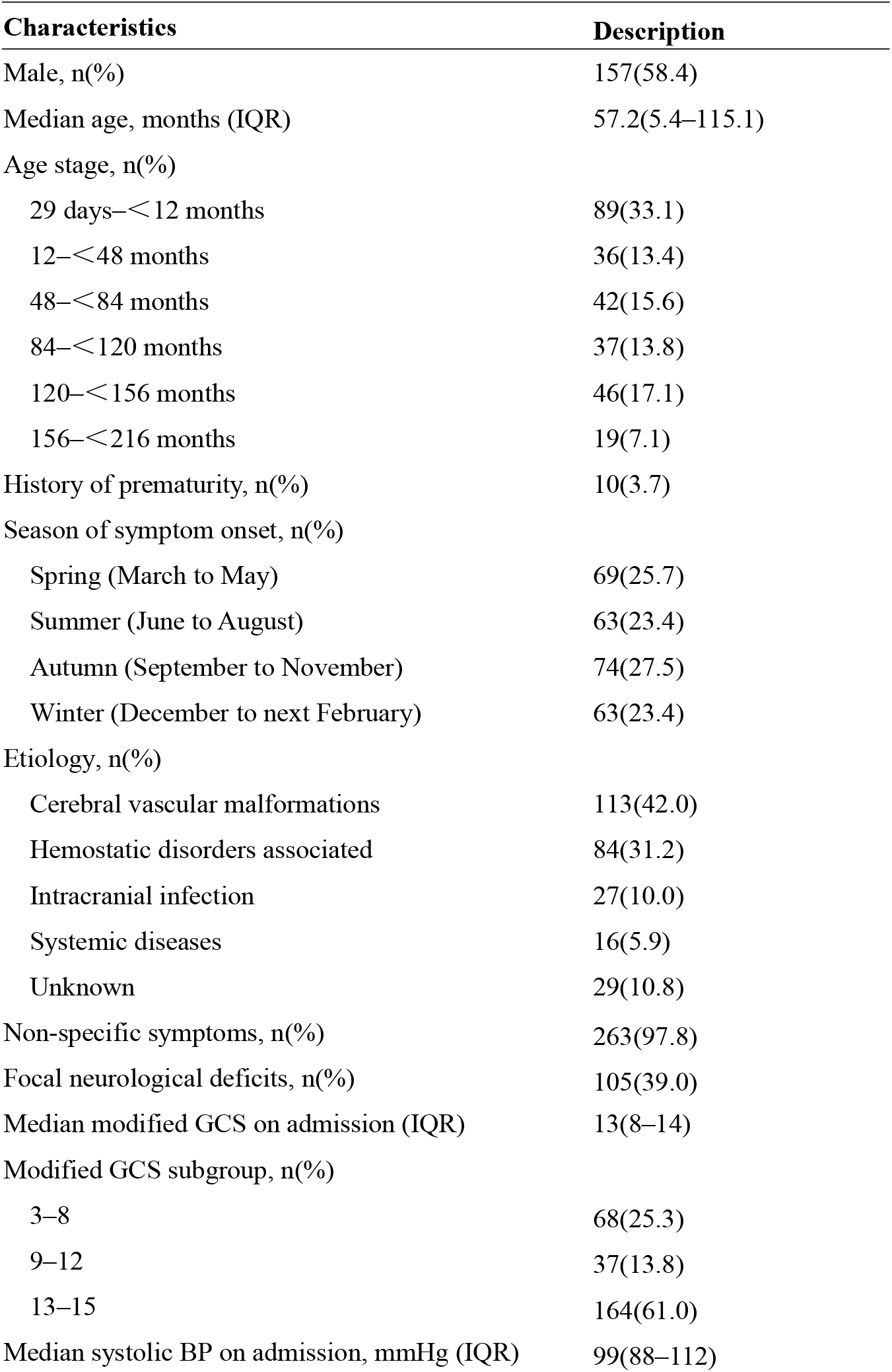
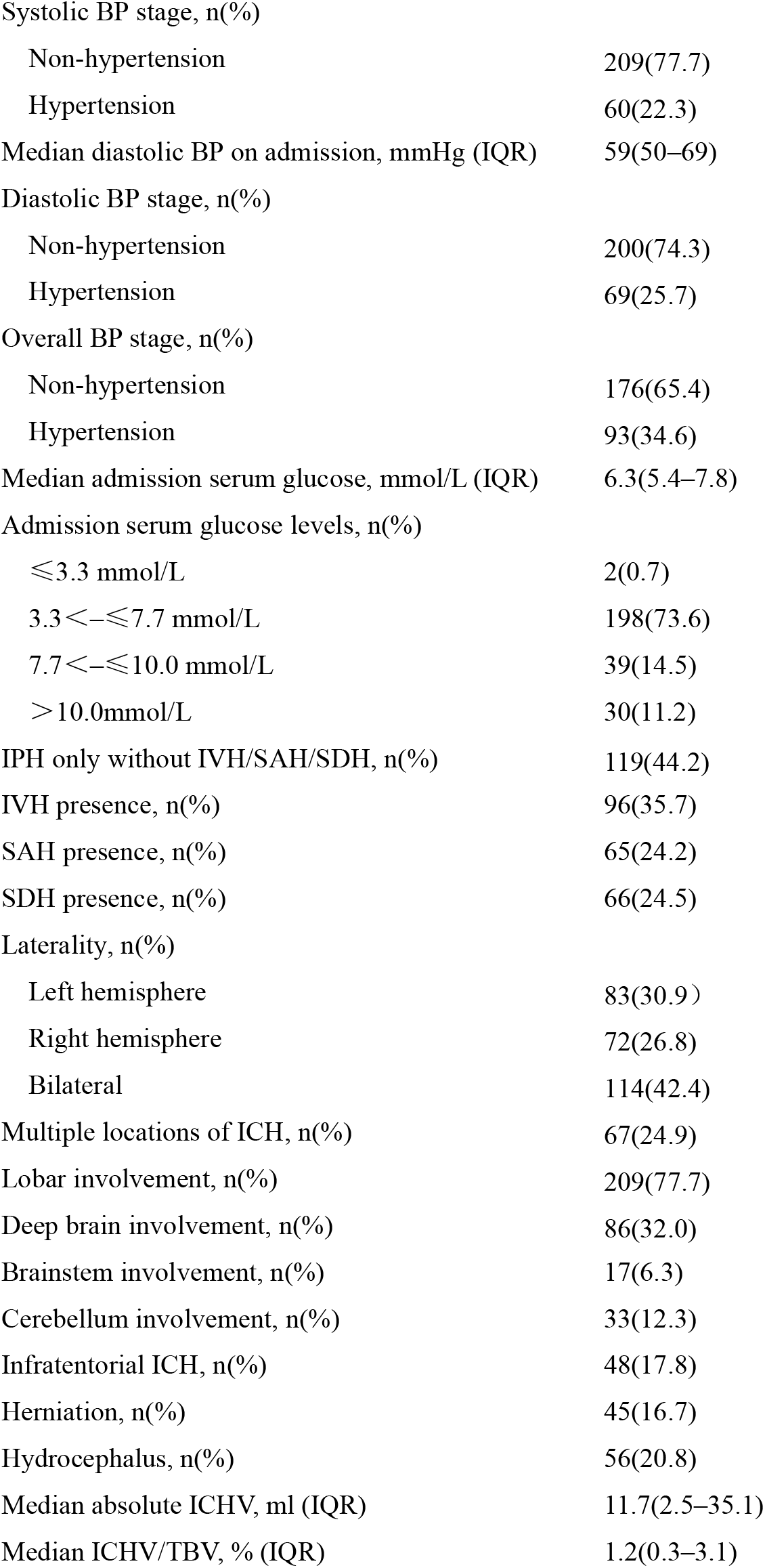
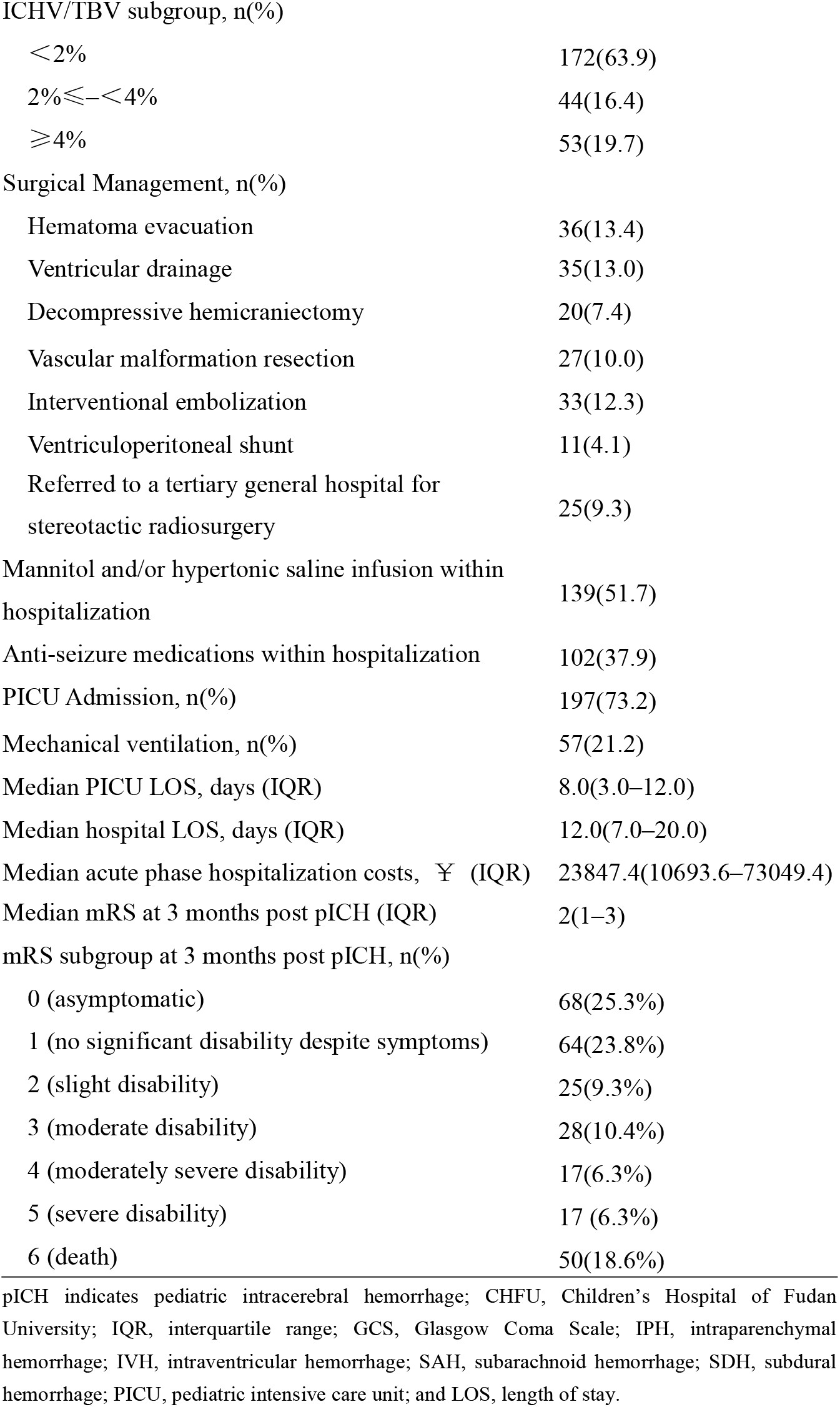
Demographics, Clinical and Hemorrhage Characteristics of CHFU pICH Cohort (n=269)

| Characteristics | Description |
| --- | --- |
| Male, n(%) | 157(58.4) |
| Median age, months (IQR) | 57.2(5.4–115.1) |
| Age stage, n(%) |  |
| 29 days–<12 months | 89(33.1) |
| 12–<48 months | 36(13.4) |
| 48–<84 months | 42(15.6) |
| 84–<120 months | 37(13.8) |
| 120–<156 months | 46(17.1) |
| 156–<216 months | 19(7.1) |
| History of prematurity, n(%) | 10(3.7) |
| Season of symptom onset, n(%) |  |
| Spring (March to May) | 69(25.7) |
| Summer (June to August) | 63(23.4) |
| Autumn (September to November) | 74(27.5) |
| Winter (December to next February) | 63(23.4) |
| Etiology, n(%) |  |
| Cerebral vascular malformations | 113(42.0) |
| Hemostatic disorders associated | 84(31.2) |
| Intracranial infection | 27(10.0) |
| Systemic diseases | 16(5.9) |
| Unknown | 29(10.8) |
| Non-specific symptoms, n(%) | 263(97.8) |
| Focal neurological deficits, n(%) | 105(39.0) |
| Median modified GCS on admission (IQR) | 13(8–14) |
| Modified GCS subgroup, n(%) |  |
| 3–8 | 68(25.3) |
| 9–12 | 37(13.8) |
| 13–15 | 164(61.0) |
| Median systolic BP on admission, mmHg (IQR) | 99(88–112) |
| Systolic BP stage, n(%) |  |
| Non-hypertension | 209(77.7) |
| Hypertension | 60(22.3) |
| Median diastolic BP on admission, mmHg (IQR) | 59(50–69) |
| Diastolic BP stage, n(%) |  |
| Non-hypertension | 200(74.3) |
| Hypertension | 69(25.7) |
| Overall BP stage, n(%) |  |
| Non-hypertension | 176(65.4) |
| Hypertension | 93(34.6) |
| Median admission serum glucose, mmol/L (IQR) | 6.3(5.4–7.8) |
| Admission serum glucose levels, n(%) |  |
| $\leq 3.3$ mmol/L | 2(0.7) |
| $3.3 < - \leq 7.7$ mmol/L | 198(73.6) |
| $7.7 < - \leq 10.0$ mmol/L | 39(14.5) |
| $> 10.0$ mmol/L | 30(11.2) |
| IPH only without IVH/SAH/SDH, n(%) | 119(44.2) |
| IVH presence, n(%) | 96(35.7) |
| SAH presence, n(%) | 65(24.2) |
| SDH presence, n(%) | 66(24.5) |
| Laterality, n(%) |  |
| Left hemisphere | 83(30.9) |
| Right hemisphere | 72(26.8) |
| Bilateral | 114(42.4) |
| Multiple locations of ICH, n(%) | 67(24.9) |
| Lobar involvement, n(%) | 209(77.7) |
| Deep brain involvement, n(%) | 86(32.0) |
| Brainstem involvement, n(%) | 17(6.3) |
| Cerebellum involvement, n(%) | 33(12.3) |
| Infratentorial ICH, n(%) | 48(17.8) |
| Herniation, n(%) | 45(16.7) |
| Hydrocephalus, n(%) | 56(20.8) |
| Median absolute ICHV, ml (IQR) | 11.7(2.5–35.1) |
| Median ICHV/TBV, % (IQR) | 1.2(0.3–3.1) |
| ICHV/TBV subgroup, n(%) |  |
| <2% | 172(63.9) |
| 2%≤–<4% | 44(16.4) |
| ≥4% | 53(19.7) |
| Surgical Management, n(%) |  |
| Hematoma evacuation | 36(13.4) |
| Ventricular drainage | 35(13.0) |
| Decompressive hemicraniectomy | 20(7.4) |
| Vascular malformation resection | 27(10.0) |
| Interventional embolization | 33(12.3) |
| Ventriculoperitoneal shunt | 11(4.1) |
| Referred to a tertiary general hospital for stereotactic radiosurgery | 25(9.3) |
| Mannitol and/or hypertonic saline infusion within hospitalization | 139(51.7) |
| Anti-seizure medications within hospitalization | 102(37.9) |
| PICU Admission, n(%) | 197(73.2) |
| Mechanical ventilation, n(%) | 57(21.2) |
| Median PICU LOS, days (IQR) | 8.0(3.0–12.0) |
| Median hospital LOS, days (IQR) | 12.0(7.0–20.0) |
| Median acute phase hospitalization costs, ¥ (IQR) | 23847.4(10693.6–73049.4) |
| Median mRS at 3 months post pICH (IQR) | 2(1–3) |
| mRS subgroup at 3 months post pICH, n(%) |  |
| 0 (asymptomatic) | 68(25.3%) |
| 1 (no significant disability despite symptoms) | 64(23.8%) |
| 2 (slight disability) | 25(9.3%) |
| 3 (moderate disability) | 28(10.4%) |
| 4 (moderately severe disability) | 17(6.3%) |
| 5 (severe disability) | 17 (6.3%) |
| 6 (death) | 50(18.6%) |

pICH indicates pediatric intracerebral hemorrhage; CHFU, Children's Hospital of Fudan University; IQR, interquartile range; GCS, Glasgow Coma Scale; IPH, intraparenchymal hemorrhage; IVH, intraventricular hemorrhage; SAH, subarachnoid hemorrhage; SDH, subdural hemorrhage; PICU, pediatric intensive care unit; and LOS, length of stay.

### Outcomes

Case-fatality was 18.2% (49/269) and 18.6% (50/269) at hospital discharge and at 3 months post pICH, respectively, all these mortalities died of the hemorrhage.

Of 220 cases surviving to hospital discharge, 110 (50.0%) had functional deficits, including cognitive impairment (92, 41.8%), sensorimotor deficits (91, 41.4%), symptomatic epilepsy (33, 15.0%) and dysphasia (23, 10.5%). All survivors had completed short-term neurological follow-up, three cases experienced re-hemorrhage (all with cerebral vascular malformations) within the short-term follow-up interval.

At a median follow-up time of 3.2 (IQR, 2.8–3.6) months post pICH, median mRS of 269 pICH was 2 (IQR, 1–3). 157 (58.4%) and 112 (41.6%) cases of 269 pICH had favorable (mRS 0–2) and unfavorable functional outcomes (mRS 3–6), respectively. Only 68 (25.3%) of 269 pICH had complete recovery with no residual symptom; 87 (39.7%) of 219 survivors had functional deficits (mRS 2–5), of whom 25 (11.4%) were slight, yet 62 (28.3%) were moderate to severe (see Table 1). Of note, functional outcome varied apparently by etiology, 81 (71.7%) of 113 cases with cerebral vascular malformation had favorable functional outcomes, conversely, a lower case-fatality (7/113=6.2%) than those without vascular malformation. Detailed distribution of mRS scores with etiology subcategory analysis are presented in Figure 2.

**Figure 2.**
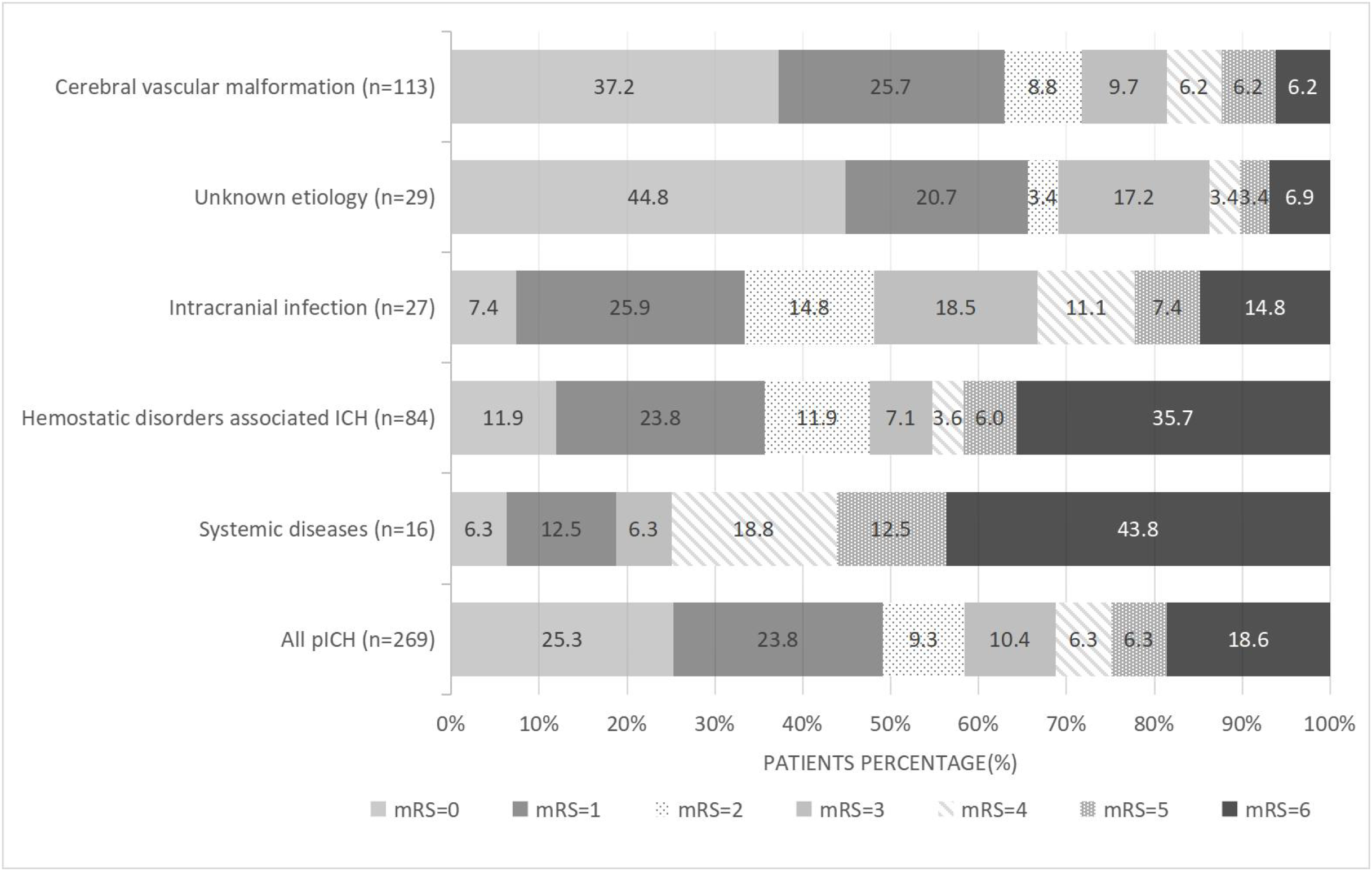
Distribution of modified Rankin Scale (mRS) scores according to etiology subcategories.

### Functional Outcome Predictors and Model Exploration

Univariate and multivariate logistic regression analyses for screening functional prognosis predictors were shown in Table 2. Multivariate analysis identified only four variables that were significantly associated with 3-month unfavorable functional outcome post pICH and composed a mnemonic code--MGBP: cerebral vascular *m*alformation as etiology (OR, 0.351[95% CI, 0.186–0.660]), modified *G*CS on admission (OR, 0.735[95% CI, 0.669–0.807]), *b*rainstem location (OR, 4.985[95% CI, 1.257–19.771]) and ICHV as *p*ercentage of TBV (OR, 1.208[95% CI, 1.059–1.377]). According to OR and 95% CI, cerebral vascular malformation and modified GCS were identified as protective factors, patients with cerebral vascular malformation were associated with a 64.9% lower risk of unfavorable functional outcome compared with those without; for per 1-point increase in GCS, the probability of unfavorable outcome decreased by 26.5%; however, brainstem location and ICHV/TBV were identified as risk factors, brainstem ICH was associated with a higher risk of unfavorable outcome compared with that non-brainstem; for per 1-percentage increase in ICHV/TBV, the probability of unfavorable outcome increased by 20.8%. Based on the regression equation, a multivariate binary logistic regression model was established to predict unfavorable functional outcome:

**Table 2.** Univariate and Multivariate Logistic Regression Analyses of Predictors for 3-month Unfavorable Functional Outcome of pICH.

| Variable | 3-month Functional Outcome of pICH<br>(n=269) |  | Univariate analysis |  |  | Multivariate analysis |  |  |
| --- | --- | --- | --- | --- | --- | --- | --- | --- |
|  | Favorable<br>outcome<br>(mRS 0–2)<br>(n=157, %) | Unfavorable<br>outcome<br>(mRS 3–6)<br>(n=112, %) | OR | 95%CI | P-value | OR | 95%CI | P-value |
| Sex |  |  |  |  |  |  |  |  |
| Male | 94(59.9) | 63(56.3) | 1.0 |  |  |  |  |  |
| Female | 63(40.1) | 49(43.8) | 1.160 | 0.710–1.896 | 0.553 |  |  |  |
| Median age, months<br>(IQR) | 61.5(3.7–114.4) | 52.0(7.1–118.0) | 0.999 | 0.995–1.004 | 0.759 |  |  |  |
| History of prematurity | 4(2.5) | 6(5.4) | 2.165 | 0.596–7.859 | 0.240 |  |  |  |
| Season of onset |  |  |  |  |  |  |  |  |
| Spring | 41(26.1) | 28(25.0) | 1.0 |  |  |  |  |  |
| Summer | 38(24.2) | 25(22.3) | 0.963 | 0.480–1.934 | 0.916 |  |  |  |
| Autumn | 44(28.0) | 30(26.8) | 0.998 | 0.512–1.947 | 0.996 |  |  |  |
| Winter | 34(21.7) | 29(25.9) | 1.249 | 0.626–2.490 | 0.528 |  |  |  |
| Cerebral vascular<br>malformation | 81(51.6) | 32(28.6) | 0.375 | 0.224–0.629 | **<0.001** | 0.351 | 0.186–0.660 | **0.001** |
| Hemostatic disorders<br>associated ICH | 40(25.5) | 44(39.3) | 1.893 | 1.123–3.190 | **0.017** | 0.811 | 0.349–1.887 | 0.627 |
| Intracranial infection | 13(8.3) | 14(12.5) | 1.582 | 0.713–3.512 | 0.259 |  |  |  |
| Systemic diseases | 3(1.9) | 13(11.6) | 6.741 | 1.873–24.257 | **0.003** | 2.989 | 0.650–13.751 | 0.160 |
| Unknown etiology | 20(12.7) | 9(8.0) | 0.527 | 0.223–1.243 | 0.144 |  |  |  |
| Median modified GCS on admission (IQR) | 14(13–15) | 9(5–13) | 0.713 | 0.653–0.780 | **<0.001** | 0.735 | 0.669–0.807 | **<0.001** |
| BP on admission |  |  |  |  |  |  |  |  |
| Non-hypertension | 111(70.7) | 65(58.0) | 1.0 |  |  | 1.0 |  |  |
| Hypertension | 46(29.3) | 47(42.0) | 1.745 | 1.049–2.902 | **0.032** | 1.374 | 0.719–2.628 | 0.336 |
| Blood Glucose on admission |  |  |  |  |  |  |  |  |
| ≤10mmol/L | 153(97.5) | 86(76.8) | 1.0 |  |  | 1.0 |  |  |
| >10mmol/L | 4(2.5) | 26(23.2) | 11.564 | 3.906–34.233 | **<0.001** | 2.913 | 0.826–10.275 | 0.097 |
| Lobe involvement | 124(79.0) | 85(75.9) | 0.838 | 0.470–1.494 | 0.549 |  |  |  |
| Deep brain involvement | 43(27.4) | 43(38.4) | 1.652 | 0.984–2.773 | **0.057** | 1.516 | 0.676–3.399 | 0.312 |
| Brainstem involvement | 5(3.2) | 12(10.7) | 3.648 | 1.247–10.671 | **0.018** | 4.985 | 1.257–19.771 | **0.022** |
| Cerebellum involvement | 17(10.8) | 16(14.3) | 1.373 | 0.661–2.849 | 0.395 |  |  |  |
| Infratentorial ICH | 22(14.0) | 26(23.2) | 1.855 | 0.989–3.479 | **0.054** | 0.729 | 0.241–2.207 | 0.576 |
| IVH presence | 49(31.2) | 47(42.0) | 1.594 | 0.962–2.640 | **0.070** | 0.701 | 0.344–1.428 | 0.328 |
| Herniation | 8(5.1) | 37(33.0) | 9.188 | 4.075–20.717 | **<0.001** | 1.780 | 0.606–5.224 | 0.294 |
| Hydrocephalus | 21(13.4) | 35(31.3) | 2.944 | 1.601–5.412 | **0.001** | 1.401 | 0.567–3.463 | 0.465 |
| Median ICHV/TBV, % (IQR) | 0.9(0.3–2.2) | 1.8(0.3–5.1) | 1.283 | 1.147–1.434 | **<0.001** | 1.208 | 1.059–1.377 | **0.005** |
| Hematoma evacuation | 16(10.2) | 20(17.9) | 1.916 | 0.944–3.889 | **0.072** | 1.224 | 0.459–3.260 | 0.686 |
| Ventricular drainage | 15(9.6) | 20(17.9) | 2.058 | 1.003–4.224 | **0.049** | 1.734 | 0.603–4.983 | 0.307 |
| Decompressive hemicraniectomy | 7(4.5) | 13(11.6) | 2.814 | 1.085–7.300 | **0.033** | 0.925 | 0.186–4.601 | 0.925 |

pICH indicates pediatric intracerebral hemorrhage; IQR, interquartile range; GCS, Glasgow Coma Scale; IVH, intraventricular hemorrhage; ICHV, intracerebral hemorrhage volume; and TBV, total brain volume.

### Logit (*P*)=3.167–1.047*cerebral vascular malformation as etiology–0.308*modified GCS on admission+1.606*brainstem location+0.189*ICHV/TBV

Receiver Operating Characteristic (ROC) analysis indicated Area Under Curve of ROC (AUC-ROC) =0.827 (95% CI, 0.778–0.877; *P*<0.001).

### Nomogram Development, Visualization and Validation

Consequently, a nomogram was developed to predict quantitatively the risk probability of unfavorable functional outcome, comprising four variables (abbreviated to MGBP) based on the visualization of the aforementioned logistic regression model. For individual patient, a higher total point of the nomogram indicated a higher probability of unfavorable functional outcome. The MGBP nomogram model and an example for using it to predict individual risk probability of 3-month unfavorable functional outcome post pICH are shown in Figure 3.

**Figure 3.**
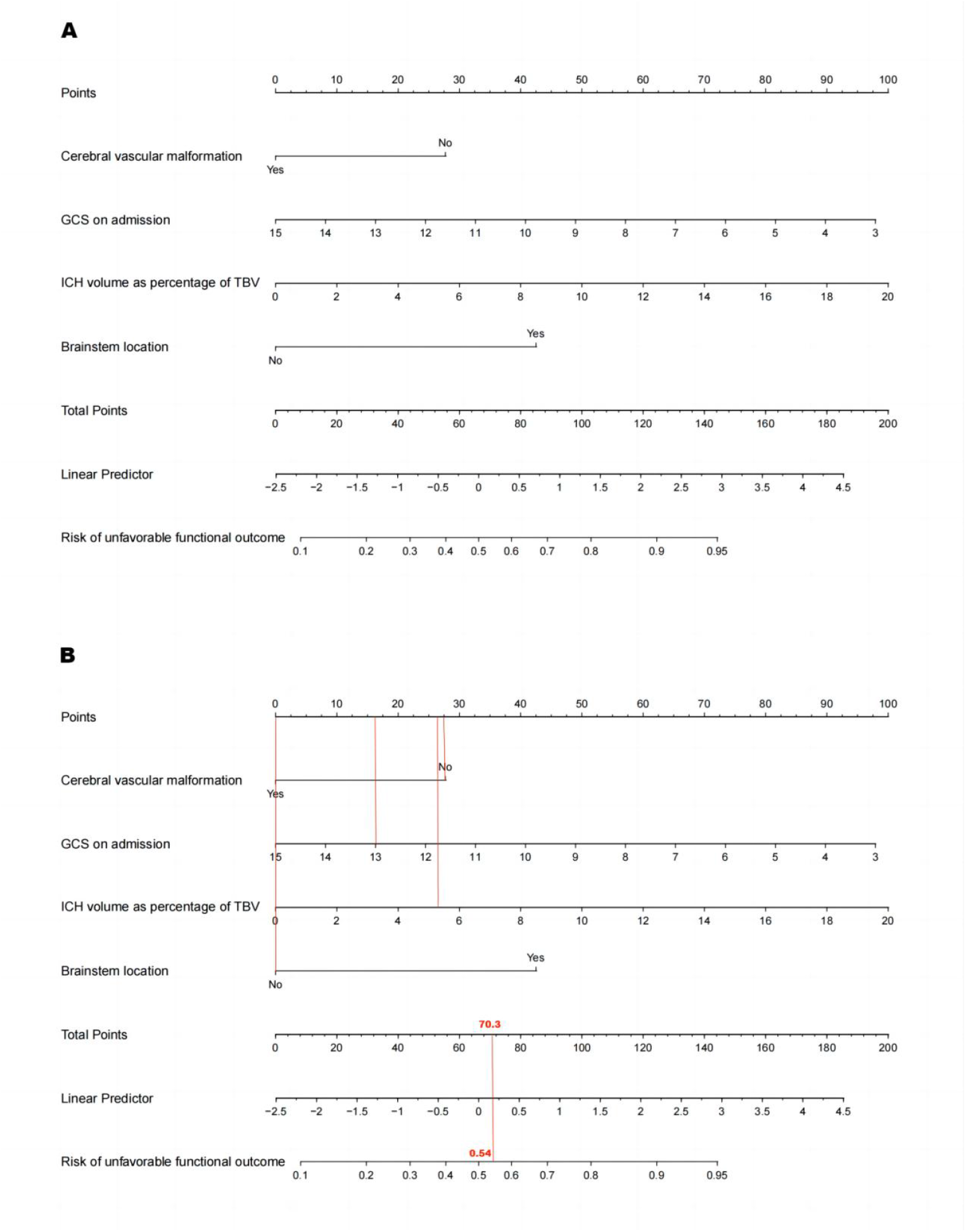
The MGBP Nomogram model. **A**, The MGBP Nomogram for prediction of unfavorable 3-month functional outcome of pediatric ICH. **B**, An example for using the nomogram, a girl with intracranial infection, modified GCS on admission of 13, lobar hemorrhage, ICH volume as 5.31% of TBV has 70.3 as total points score according to this graphic calculation tool and are converted into an estimated individual probability of 3-month unfavorable functional outcome post pediatric ICH of 54%. GCS indicates Glasgow Coma Scale; ICH, intracerebral hemorrhage; and TBV, total brain volume.

The ROC curve (Figure 4A) of the MGBP nomogram with C-Statistics=0.827 reflected a favorable discrimination. Moreover, the nomogram was internally validated using bootstrap, the *P*-value=0.803 for the Spiegelhalter’s Z-test indicated no significant departure from a well fit, the Brier score=0.163 suggested the calibration curve (Figure 4B) was almost close to the ideal curve, revealed a high consistency of the nomogram. Additionally, the Decision curve analysis (Figure 4C) showed a wide range of risk thresholds and considerable net benefits in the nomogram model than in the All or None scheme if the threshold probability was between 0.1 and 0.9. All these favorable model performance measurements demonstrated that via bridging the gap between prognostic factors exploration and clinical application, the MGBP nomogram had a potential utility for functional prognosis prediction, clinical decision making and stratified analysis of clinical research.

**Figure 4.**
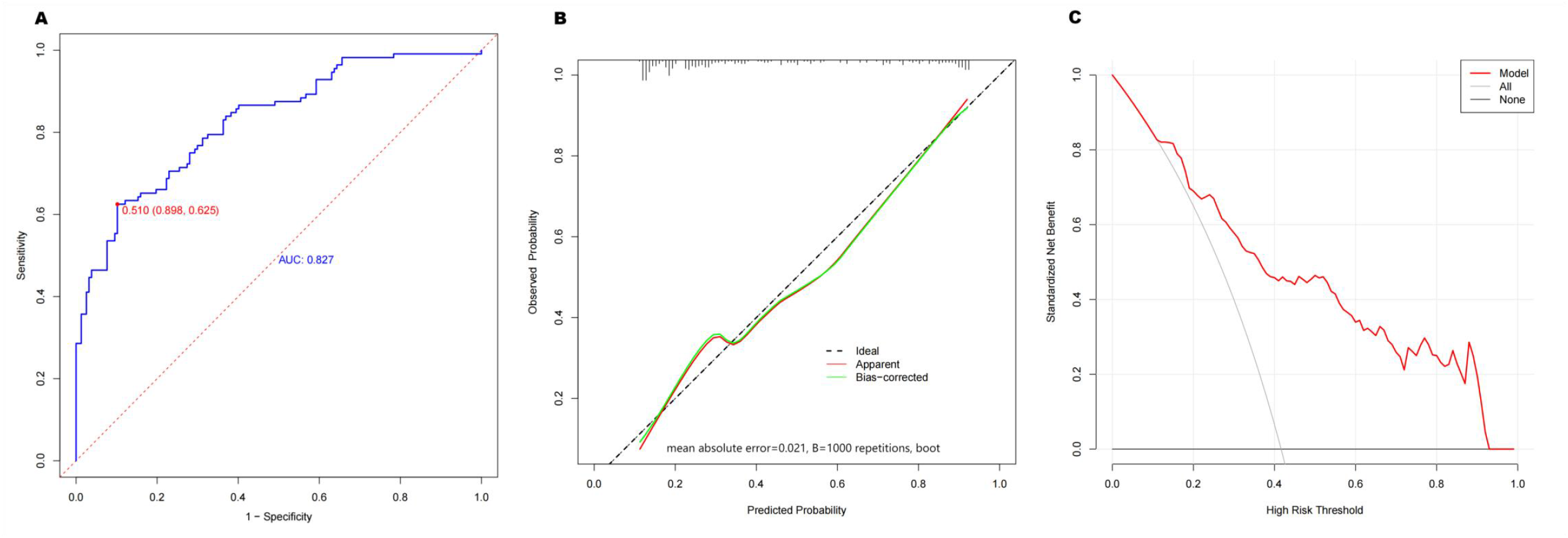
Performance measurements of the MGBP nomogram model. **A**, Receiver operating characteristic curve for prediction probability of unfavorable functional outcome of pediatric intracerebral hemorrhage (pICH). **B**, Calibration curve for prediction probability of unfavorable functional outcome of pICH. **C**, Decision curve analysis for prediction probability of unfavorable functional outcome of pICH.

## Discussion

### Implications

This single-center retrospective study described detailed features of nontraumatic pICH in a hospital-based consecutive cohort, with the largest non-neonate pICH cohort to date, we performed multivariate analysis to screen more than 10 candidate functional prognosis predictors in a comprehensive way; thus, based on four clinical and radiological predictors (cerebral vascular *m*alformation as etiology, modified *G*CS on admission, *b*rainstem location and ICHV as *p*ercentage of TBV), the MGBP is the first nomogram model developed and validated specifically in a Chinese pICH cohort for individual quantitative unfavorable functional outcome prediction.

Univariate and multivariate analyses concluded that sex, age, HDICH, intracranial infection, systemic diseases as etiology, unknown etiology, hypertension, hyperglycemia, lobe, deep brain, cerebellum location, infratentorial ICH, IVH presence, herniation syndrome, hydrocephalus, hematoma evacuation, ventricular drainage, decompressive hemicraniectomy were not significant predictors of unfavorable functional outcome, it did not imply that these variables were completely uncorrelated with functional outcome, however, their effects reflected via four powerful and easily available predictors (all components of the MGBP nomogram) which were significantly associated with functional prognosis, thus, the nomogram would benefit from simplifying prediction model, and this practical and reliable approach could be assessed by clinicians rapidly in the acute settings for risk stratification and to tailor further aggressive management as to save public health resources.

Two prognostic scores were proposed in previous studies to predict outcome post pICH, the pediatric ICH (PICH) score (four components including IPH volume as percentage of TBV, hydrocephalus, herniation and infratentorial, ranging 0–5) created by Beslow et al^30^ and the modified PICH (mPICH) score (six components consisting of herniation, initial altered mental status, hydrocephalus, infratentorial, IVH and ICHV/TBV > 2%, ranging 0–13) constructed by Guédon et al,^8^ using King’s Outcome Scale for Childhood Head Injury score as outcome measurement, both scores were integer-based point scoring systems for each variable, the total points were calculated as sum of the variable weighted points; comparatively, the MGBP nomogram model was developed base on mRS as outcome measurement, making full use of variable information, the total points of the nomogram were converted into a continuum of individual quantitative risk probability through logarithmic formula as to improve predictive accuracy of the model.

Additionally, we included an etiology indicator in the MGBP nomogram as cases with vascular malformation as etiology in our cohort had better functional outcomes and the lowest case-fatality compared with those of cases with other etiology categories, it was similar to data published from a cohort study which indicated that children with vascular lesions as etiology of pICH had significantly better clinical functional outcomes and a lower mortality than those with other etiologies.^5^ Furthermore, our multivariate analysis corroborated that vascular malformation as etiology was a predictor of functional outcome, it might be partially explained by the limitation of cerebral vascular malformation impairment within the central nervous system and without other organ dysfunction, therefore, acute and subsequent therapies are relative specific.

Different from PICH and mPICH scores which has no assessment and qualitative assessment of mental status respectively, the MGBP nomogram included a clinical indicator, modified GCS on admission, as discrete variable to make full use of information and to assess mental status quantitatively and more accurately, multivariate analysis also confirmed GCS as an important functional prognosis predictor and lower GCS to be associated with increased risk of unfavorable outcome, which was consistent with a previous study in PICU from Italy reported that a low GCS on PICU admission was predictive of severe neurological impairment in survivors with sICH at short-term follow-up,^31^ suggesting on admission GCS may serve as a hyperacute warning indicator of functional outcome, and it could be obtained within a few minutes after hospitalization through bedside assessment which highlights its convenience.

Multivariate analysis corroborated that brainstem location was a predictor of functional outcome, nevertheless, neither cerebellum location nor infratentorial ICH was significantly associated with functional outcome, indicating the importance of identifying brainstem ICH from all infratentorial ICH. Presence of IVH did not predict functional outcome neither, which was consistent with prior investigations.^31-33^

Multivariate analysis concluded ICHV/TBV as a robust functional prognosis predictor, this was in keeping with published literature^28,34^ which showed large percentage of ICHV/TBV predicted poor neurological outcome. Through relatively non-invasive simple measurements, clinicians could estimate ICHV/TBV in less than a few minutes at the early stage of clinical practice, relatively objective quantified assessment was another highlight of this indicator. Furthermore, compared with the risk-grouping categorizations for percentage of ICHV/TBV used by PICH and mPICH scores (< 2%, 2–3.99%, > 4%, and > 2%, respectively), we revealed that ICHV/TBV as a continuous variable was the strongest predictor of functional prognosis and had the largest weight coefficient in the MGBP nomogram, and thereby the predictive accuracy had been improved.

Furthermore, our findings regarding prevalence of cerebral vascular malformation as etiology of pICH (42.0%) and, among which, of AVM, were partly similar to a recent meta-analysis of 1309 patients across 23 studies, which reported that the dominant etiology of pICH in children after neonatal period was cerebral vascular lesion (aggregate prevalence of 59%, more frequent in children 2–18 years old), followed by hematological disorders (12%), and AVM represented the most frequent cause (68.3% of all vascular lesions).^35^ However, in our cohort, HDICH (among which idiopathic VKDB representd for 34.5%) accounted for a greater proportion of pICH (31.2%) than in the foregoing literature and previous studies from North America or Europe, wherein HDICH was mainly caused by hemophilia which accounted for approximately 90% of all inherited coagulation disorders.^36,37^ Additionally, a single-center case-control study from China reported cerebrovascular disease (37.0%) and hematologic system disease (33.5%) were the most frequent categories of pICH, AVM (28.5%) and VKDB (20.5%) were the specific leading causes of pICH, these were quite consistent with our results.^14^ A major driver of this regional disparity is the higher prevalence of vitamin K deficiency in Chinese infants than in western populations, as Chinese infants were usually breastfed only during their first year of life, and large proportion of whom without prophylactic vitamin K administration after birth, prior case-series reports from China also advocated the prevalence of VKDB in pediatric HS or ICH up to over 50%.^10,11^ Furthermore, 10% pICH remained idiopathic in this study, which was consistent with a recent review demonstrated 14% patients had pICH of unknown etiology,^36^ indicating the importance of comprehensive assessment and long-term follow-up.

Prior randomized clinical trials showed early intensive BP lowering (EIBPL) to a target of SBP<140 mmHg was not associated with a lower rate of death or disability at 3 months post ICH in adults;^38,39^ meanwhile, hyperglycemia on admission was strongly associated with death or disability at 3 months post ICH in adults.^40,41^ However, a significant knowledge gap exists regarding the relationship between hypertension, hyperglycemia and functional outcome in children with pICH. In our study population, hypertension (only present in one-third cases of pICH) was not prevalent, whilst hyperglycemia (only present in one-tenth cases) was relatively not common; in multivariate analysis, neither hypertension nor hyperglycemia was significantly associated with unfavorable functional outcome, although hyperglycemia showed a trend (*P*=0.097) toward unfavorable outcome; yet both of them were associated with unfavorable functional outcome in our univariate analysis. Our research was consistent with a retrospective case-control study which reported that in childhood AIS, hypertension had no relationship with neurological poor outcome or death;^42^ nevertheless, still in this study indicated that blood glucose level≥11.1mmol/L was associated with poor neurological outcome, which differed from our study. Our findings could be explained via the following possibilities: first, protective autoregulation mechanism driven by our body which increases BP to elevate intracranial pressure to maintain adequate cerebral perfusion and minimize secondary brain impairment; additionally, the pathogenesis of pICH is often different from that of adult ICH, cerebral vascular malformation is the main etiology of pICH, meanwhile adults tend to have hypertensive encephalopathy more; furthermore, inadequate pediatric hypertension awareness both in society generally and among first-line healthcare providers as well as absence of regular basic screening, pediatric hypertension prior to pICH have potential risk of being underdiagnosed.

2022 guideline of sICH recommend that in adult sICH of mild to moderate severity with SBP 150–220 mmHg, acute lowering of SBP to 140 (target range of 130–150) mmHg is safe and reasonable to improve functional outcomes; and treating moderate to severe hyperglycemia (> 10.0–11.1 mmol/L) in adults with sICH is reasonable to improve outcomes.^1^ Given the lack of adequate pediatric data, optimal BP and glucose managements in pICH have not been defined, the role of EIBPL and intensive glucose control are unknown, further clinical trials deserve investigating to determine optimal thresholds and target ranges of these potentially modifiable physiological parameters in pICH.

High critical care utilization (73.2% PICU admission) of pICH in our study was similar to a population-based study from the US which reported 73% of children with HS were admitted to PICU.^6^ Case-fatality rates at hospital discharge (18.2%) and at 3 months post pICH (18.6%) were similar to published data from China (16.5% at hospital discharge)^14^ and from a recent meta-analysis (17.3% at last follow-up).^5^ In terms of functional outcome, 50% and 39.7% survivors of pICH had functional deficits at hospital discharge and at 3 months post pICH, respectively, which would lead to heavy burdens and this was consistent with published literature which reported that approximately 40–70% survivors had functional deficits.^5-7,12,15,43,44^

### Limitations

There are some limitations in this study. First, owing to the extremely low incidence of pICH, it was a single-center retrospective design study, a national children’s medical center-based cohort rather than a population-based sample that may have a potential for Berkson’s bias. As an exploratory study for developing a predictive nomogram, we only performed internal validation, future external validation and extrapolation feasibility evaluation in a multicenter, prospective cohort are required to further elucidate generalizability of the MGBP nomogram. Second, as considering tumor itself might affect functional prognosis, we excluded patients with intracranial tumor which may result in exclusive bias, although the proportion accounted for only a small part (11/298=3.7%) and was similar to data published from a recent review (4%),^36^ future studies with more detailed assessments on this subgroup are needed.

Third, hypertension and hyperglycemia were defined based on admission measurements, further randomized clinical trials with multiple measurements at various time points (prehospital, on admission, 24, 48 and 72 hours) and variability deserve investigating to evaluate the effects of persistent hypertension and hyperglycemia on mortality or functional outcome of pICH.

Finally, at our institution, not all subcategories of pICH would complete long-term follow-up which extends years especially at an early stage in the past decade, future research with meaningful long-term assessments needs to be well characterized as to evaluate the development of residual functional deficits with systemic rehabilitation, additionally, potential emotional concerns, academic plus adaptive functioning impairment may appear over time since pediatric survivors grow up.

## Conclusions

The MGBP nomogram integrating four robust predictors together (cerebral vascular *m*alformation as etiology, modified *G*CS on admission, *b*rainstem location and ICHV as *p*ercentage of TBV) as a novel and potentially effective approach could reliably predict individual quantitative risk probability of 3-month unfavorable functional prognosis post pICH, thereby yielding a bridge between clinical and radiological features and personalized precision medicine within clinical practice.

## Data Availability

The data sets that support the findings of this study are available from the corresponding author on reasonable request.

## Nonstandard Abbreviations and Acronyms

CHFU: Children’s Hospital of Fudan University
GCS: Glasgow Coma Scale
HDICH: Hemostatic disorders associated intracerebral hemorrhage
ICH: intracerebral hemorrhage
ICHV: intracerebral hemorrhage volume
mRS: modified Rankin Scale
pICH: pediatric intracerebral hemorrhage
TBV: total brain volume

## Acknowledgements

The authors thank Dr. Zhu-Jin Lu for his helpful comments in revising the manuscript.

## Supplemental Material

Tables S1–S4

Figures S1–S2

TRIPOD Checklist

